# Lessons from movement ecology for the return to work: modeling contacts and the spread of COVID-19

**DOI:** 10.1101/2020.05.27.20114728

**Authors:** Allison K. Shaw, Lauren A. White, Matthew Michalska-Smith, Elizabeth T. Borer, Meggan E. Craft, Eric W. Seabloom, Emilie Snell-Rood, Michael Travisano

**Affiliations:** Department of Ecology, Evolution, and Behavior, University of Minnesota, St. Paul, Minnesota, USA; National Socio-Environmental Synthesis Center, Annapolis, Maryland, USA; Department of Veterinary Population Medicine, University of Minnesota, St. Paul, Minnesota, USA; Department of Plant Pathology, University of Minnesota, St. Paul, Minnesota, USA; BioTechnology Institute, University of Minnesota, St. Paul, Minnesota, USA

**Keywords:** contact network epidemiology, disease ecology, infection, mathematical model, network, SARS-CoV-2, theoretical ecology, theory, transmission

## Abstract

Human behavior (movement, social contacts) plays a central role in the spread of pathogens like SARS- CoV-2. The rapid spread of SARS-CoV-2 was driven by global human movement, and initial lockdown measures aimed to localize movement and contact in order to slow spread. Thus, movement and contact patterns need to be explicitly considered when making reopening decisions, especially regarding return to work. Here, as a case study, we consider the initial stages of resuming research at a large research university, using approaches from movement ecology and contact network epidemiology. First, we develop a dynamical pathogen model describing movement between home and work; we show that limiting social contact, via reduced people or reduced time in the workplace are fairly equivalent strategies to slow pathogen spread. Second, we develop a model based on spatial contact patterns within a specific office and lab building on campus; we show that restricting on-campus activities to labs (rather than labs and offices) could dramatically alter (modularize) contact network structure and thus, potentially reduce pathogen spread by providing a workplace mechanism to reduce contact. Here we argue that explicitly accounting for human movement and contact behavior in the workplace can provide additional strategies to slow pathogen spread that can be used in conjunction with ongoing public health efforts.

## Introduction

The cosmopolitan connectivity of modern society facilitated the rapid spread of SARS-CoV-2 around the globe in early 2020 [1]. The rate at which any pathogen spreads depends critically on host movement behavior [2]. Indeed, estimates of key epidemiological parameters like the basic reproduction number (R_0_) are highly variable in part because they are context-specific and are a function of behaviors like movement and heterogenous contact structure [3,4]. Although most cases of COVID-19 (the disease caused by SARS-CoV-2) seem to be mild or even asymptomatic [5,6], the sheer number of cases to date means that limited personnel, hospital beds, and ICU equipment can be rapidly overwhelmed, increasing mortality [7,8]. Thus, continuing normal movement patterns, unmitigated, is not a viable containment strategy. Without a vaccine or widespread immunity to SARS-CoV-2, our best defense to slow pathogen spread has been restricting movement and contacts through physical distancing [9], testing for SARS-CoV-2 when available [10] and contact tracing [11]. Lockdown measures have drastically reduced human movement [1,12] and consequently have reduced the effective reproduction number, R_e_ [4,13]. However, such measures are affecting mental health [14,15] and have had a devastating impact on the economy, so individual regions are considering best practices for the reopening of businesses, schools, and other places where people gather (e.g.,[16—18]). Decisions regarding next steps can be informed by recognizing that not all movement patterns nor all contact behaviors are equal in terms of pathogen spread.

Concepts from movement ecology and contact network epidemiology can provide helpful frameworks for understanding the nuanced interactions between movement, contacts and infection. Increased movement does not always mean increased transmission risk [19]; for example, movement that either takes individuals away from infected areas or reduces contact with infected conspecifics can reduce transmission risk (migratory escape; [20,21]). Increased movement can even increase some aspects of infection risk while decreasing others, simultaneously [22]. Thus, explicitly considering how movement relates to transmission can help us understand what effect different movement patterns have on infection dynamics [23,24]. Similarly, from disease ecology and contact network epidemiology, we know that structured contacts among individuals in a population have different effects on disease spread than random contacts. For example, long-range connections in otherwise locally-connected small world networks can have dramatic effects on disease spread at a population level [25].

Individual movement across multiple scales — from occasional global movements to smaller-scale daily patterns — is critical for shaping contact and thus the spread of pathogens. To date, models of SARS-CoV-2/COVID-19 spread have focused on comparing patterns of spread across countries, states, and counties [26,27]. Indeed, a plethora of epidemiological models have proven useful in generating recommendations for reducing the virus spread rate, from understanding the role of contact-tracing and society-wide physical distancing [11,28], to travel restrictions and lockdowns (e.g., [4,29,30]), to mask-wearing [31]. However, few models offer guidance at scales as fine as individual workplaces, despite the fact that this local scale is where individual decisions are made and where most transmission occurs. Furthermore, apart from time at home, the most predictable component of many people’s days is time spent in the workplace. Thus, knowledge of work commute patterns, contact networks of individuals in the workplace, and related workplace-specific factors could help mitigate pathogen spread during the period that total population immunity remains low. In many cases, commute trajectories are not random but involve regularity in timing, location, and encounters with other individuals along the way (e.g., on public transport). Here, we consider the implications for mitigating COVID-19 transmission using a case study of the initial stages of resuming research at a large research university.

Implicit in this analysis is that COVID-19 is currently spreading in local communities around the world, and every individual in a workplace is part of a home community. Even under many weeks of extreme restrictions with only society’s most essential employees present in workplaces (i.e., Stay at Home orders), the number of new cases have continued to rise in most locations. For example, in late April 2020, even after three weeks of a Stay at Home order and extreme physical distancing in Minnesota, a state with moderate spread and commendable compliance with the order, the number of new cases confirmed each day had tripled [32]. With community spread of this pathogen, it is unrealistic to expect zero workplace infection or widespread virus containment primarily through workplace practices. Any return-to-work plan, therefore, must include the explicit expectation that new infections may arise while concurrently prioritizing worker safety and optimizing the work that can be done. Thus, reopening businesses requires an evidence-based plan to reduce contacts through time to minimize new infections at the workplace, when an infected individual, presumably pre-symptomatic [33], brings the virus to a workplace.

Here we develop a pair of models to understand how movement and contact structure shape infection spread. As a case study, we consider the context of moving from full-time work at home to part-time resumption of research at a university; however, results from this model are general to many other settings as well. We take a dual modeling approach by developing a general movement model and a network case study of one academic laboratory and office building. We explore tradeoffs between limiting contact, people, or time on campus. We find that moving back to work on campus does not necessarily speed up infection spread, and depends particularly on the infection risk associated with commutes and how well physical distancing can be maintained on campus. Thus our findings allow us to set evidence-based expectations and generate specific behavioral recommendations for a safer return to work.

## Materials and Methods

We develop two models: a movement model to explore movement between home and work environments, and a network model to explore contact patterns within the work environment. Both models are SEIR, tracking susceptible (*S*), exposed *(E)*, infected *(I)*, and removed (recovered and immune, or deceased; R) individuals. We assume there is no loss of immunity (removed individuals never move back to the susceptible class) over the short time scales we consider, and we assume a closed population (no births, deaths, immigration, or emigration).

## Methods: movement model

### Setup

Our first model explores infection dynamics as individuals move between home, commuting, and work environments. Workplaces, including universities, face a number of different decisions about how to slowly ramp up work following easing of lockdown. Here, we simulate three potential strategies for returning to work: (i) allowing people to return while maintaining physical distancing, (ii) limiting the number of people returning to campus during the work day, and (iii) limiting the time each person spends on campus. For each strategy combination, we simulate infection dynamics and quantify two output metrics: (1) the ‘final epidemic size’ (cumulative fraction of the population infected, at equilibrium), and (2) the ‘epidemic peak size’ (maximum fraction of the population infected during the outbreak). The aim of this type of conceptual model is to clarify the connections between assumptions and outcomes, unlike predictive models which would contain an abundance of empirical data and aim to generate forecasts for a specific system [34].

### Daily cycle

Our model dynamics have a combination of continuous and discrete time (e.g., [35]), where each day is broken into discrete phases *(T*_h_ spent at home, *T*_w_ spent at work, and *T*_c_ spent commuting each way, with *T*_h_ + *T*_w_ + 2*T*_c_ = 1) and infection dynamics occur continuously during each phase (Fig 1, see Tables 1-2 for model variables and parameters). All individuals start at home and spend a fraction of their day (of length *T*_h_) there and not working. During this time, the infection dynamics are given by

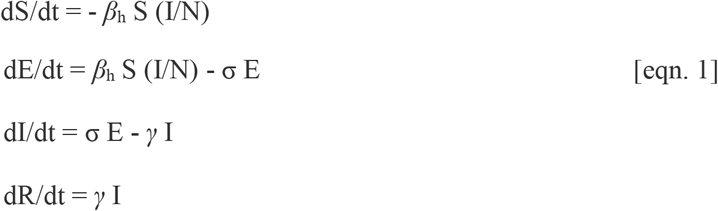

where *S* is the number of susceptible individuals, *E* is the number of exposed individuals, *I* is the number of infected individuals, *R* is the number of removed (recovered and immune, or deceased) individuals, *N* is the total number of individuals in the population (*N* = *S* + *E* + *I* + *R*), *β*_h_ is the rate of transmission while at home, σ is the rate of moving from exposed to infected (inverse of the latent period) and γ is the rate of recovery from infection.

**[Fig 1.**
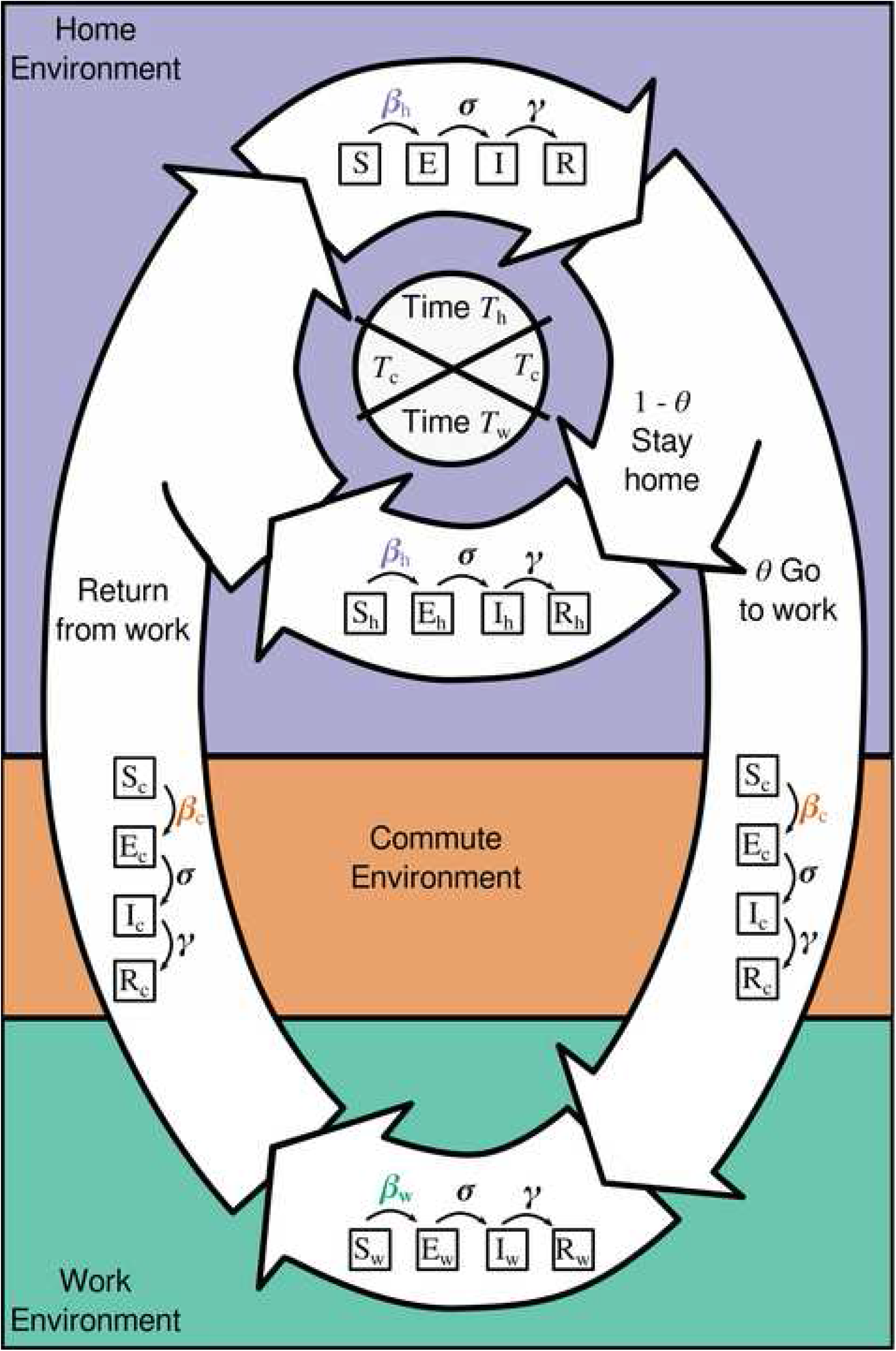
Movement model schematic, showing a daily cycle. All individuals — Susceptible (S), Exposed (E), Infected (I) or Removed (R) — spend part of their day (*T*_h_) at home. A proportion *θ* of individuals move to campus, spending *T*_c_ time commuting in each direction, and work from campus during the workday (time *T*_w_), while the other fraction (1 - *θ*) works from home. A total 24 hour cycle is then represented by: *T*_h_ + *T*_w_ + 2 *T*_c_ = 1. Transmission rates can vary among home (*β*_h_; this includes transmission during essential trips e.g., to the grocery store), commute *(β*_c_; traveling between home and work), and work (*β*_w_; campus-based interactions) environments, while the rate of moving from exposed to infected (σ) and recovery rate (γ) are the same regardless of where individuals are located.]

**Table 1.** Movement model state variables and their meaning.

| Variable | Meaning |
| --- | --- |
| $S$ | Total number of susceptible individuals |
| $E$ | Total number of exposed individuals |
| $I$ | Total number of infected individuals |
| $R$ | Total number of removed individuals |
| $S_c$ | Number of susceptible individuals commuting to campus (during the commute phases) |
| $E_c$ | Number of exposed individuals commuting to campus (during the commute phases) |
| $I_c$ | Number of infected individuals commuting to campus (during the commute phases) |
| $R_c$ | Number of removed individuals commuting to campus (during the commute phases) |
| $S_w$ | Number of susceptible individuals working from work (during the work phase) |
| $E_w$ | Number of exposed individuals working from work (during the work phase) |
| $I_w$ | Number of infected individuals working from work (during the work phase) |
| $R_w$ | Number of removed individuals working from work (during the work phase) |
| $S_h$ | Number of susceptible individuals working from home (during the commute and work phases) |
| $E_h$ | Number of exposed individuals working from home (during the commute and work phases) |
| $I_h$ | Number of infected individuals working from home (during the commute and work phases) |
| $R_h$ | Number of recovered individuals working from home (during the commute and work phases) |

**Table 2.** Movement model parameters, meaning, and default value (with units).

| Parameter | Meaning | Default values<br>[Units] | Sensitivity<br>analysis range |
| --- | --- | --- | --- |
| $N$ | Population size | 3000 [people] | (1500 to 6000) |
| $R_0$ | Basic reproductive number (number of new infections that each infection generates) | 2.5 [unitless] [39] | fixed |
| $R_{e-c}$ | Effective reproductive number while commuting between work and campus | $R_0$ [unitless] | (1 to 4 $R_0$) |
| $R_{e-h}$ | Effective reproductive number while at home | $0.5R_0$ [unitless] [40] | (0.25 $R_0$ to $R_0$) |
| $R_{e-w}$ | Effective reproductive number while at work at campus | $R_0$ [unitless] | (0.5 $R_0$ to 2 $R_0$) |
| $T_c$ | Fraction of a 24-hour day spent commuting each way for those that commute to campus | 1/24 [unitless] | (0.5/24 to 2/24) |
| $T_h$ | Fraction of a 24-hour day spent not working (everyone is off campus) | $= 1 - 2 T_c - T_w$<br>[unitless] | $= 1 - 2 T_c - T_w$ |
| $T_w$ | Fraction of a 24-hour day spent at work on campus for those commuting (some individual are on campus) | 8/24 [unitless] | (2/24 to 12/24) |
| $\beta_c$ | Transmission rate while commuting | $= \gamma R_{e-c}$ [day<sup>-1</sup>] | $= \gamma R_{e-c}$ |
| $\beta_h$ | Transmission rate while at home | $= \gamma R_{e-h} [\text{day}^{-1}]$ | $= \gamma R_{e-h}$ |
| $\beta_w$ | Transmission rate while at work | $= \gamma R_{e-w} [\text{day}^{-1}]$ | $= \gamma R_{e-w}$ |
| $\sigma$ | Rate of moving from exposed to infected (inverse of the latent period) | 1/4.2 [day<sup>-1</sup>] [37] | (1/5.1 to 1/3.5) |
| $\gamma$ | Recovery rate | 1/9.5 [day<sup>-1</sup>] [38] | (1/11 to 1/6) |
| $\theta$ | Fraction of the campus population commuting to work on campus (instead of continuing to work at home) | 1 [unitless] | (0.0001 to 1) |

Here, the rate at which new susceptible individuals (*S*) become exposed (*E*) depends on three components [36]. First is the rate of contact between two individuals in a location. Here we assume this contact rate is constant (does not change with population density) but can differ across environments (home vs. work vs. commuting). Critically, we assume that *β*_h_ accounts for transmission not just in an individual’s actual home, but transmission that occurs during other essential activities during lockdown (e.g., grocery store trips). Second is the probability that the contact for each susceptible individual is with an infected individual; this is given by the proportion of infected individuals in the local population *(I/N)*. Third is the probability that contact with an infectious individual results in transmission. In equation 1 above (and the other equations below), we have combined the first and third factors into a single term, *β*, while the second factor is given by *I*/*N*. Overall, this gives us frequency-dependent transmission (transmission rate depends on the frequency — not density — of infected individuals in the population); an appropriate assumption for spatially structured environments [2,36].

After the period of time at home (*T*_h_), a fraction, *θ*, of all individuals commute to work while the remaining (1- *θ*) stay to work from home. At this point we subdivide the population based on the number of individuals of each type and fraction commuting. We denote location by subscripts (*h* for home, *c* for commute), so the number of individuals of each type are

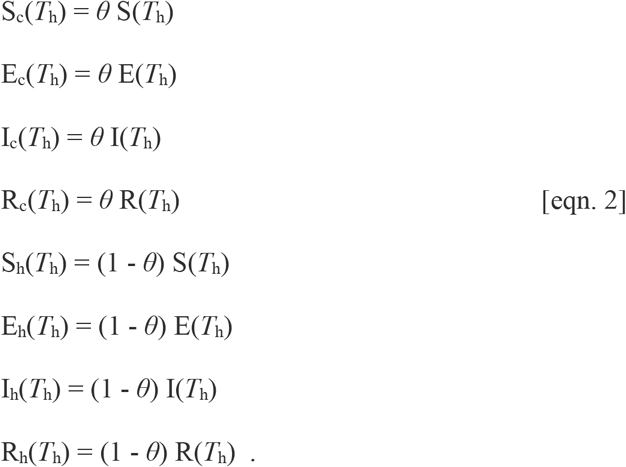

During the commute phase, the infection dynamics for those commuting are given by

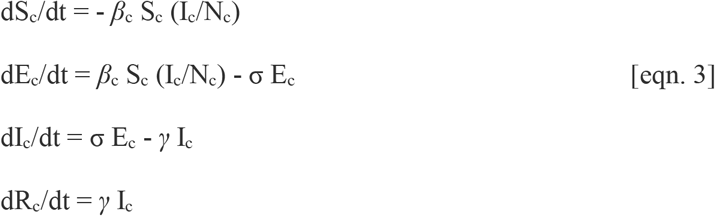

where *N*_c_ is the total number of individuals commuting (*N*_c_ = *S*_c_ + *E*_c_ + *I*_c_ + *R*_c_), and *β*_c_ is the rate of transmission while commuting. Similarly, during the commute phase, the infection dynamics for thosed

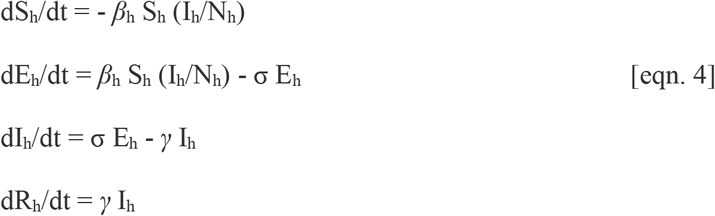

where *N*_h_ is the total number of individuals at home *(N*_h_ = *S*_h_ + *E*_h_ + *I*_h_ + *R*_h_).

After the commute phase (of length T_c_), comes a work phase. Here, the population continues to be subdivided into eight types, where the number of individuals of each type are

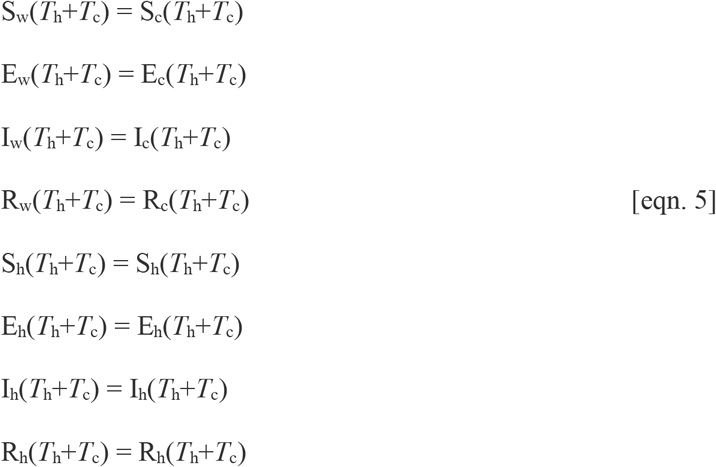

where the subscript *w* denotes work. Note that individuals that work on campus switch from a commute (c) subscript to a work (w) one here, while individuals that work at home continue with the same subscript (h). Both groups are still experiencing infection dynamics. During the work phase, the infection dynamics in the workplace are given by

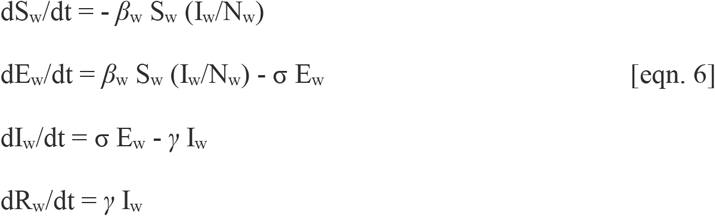

where *N*_w_ is the total number of individuals at work (*N*_w_ = *S*_w_ + *E*_w_ + *I_w_* + *R*_w_), and *β*_w_ is the rate oftransmission while at work. During the work phase, the infection dynamics for those working at home are given by [eqn. 4] above.

After the work phase (of length *T*_w_), we describe a second commute phase. The population continues to be subdivided into eight types, where the number of individuals of each type are

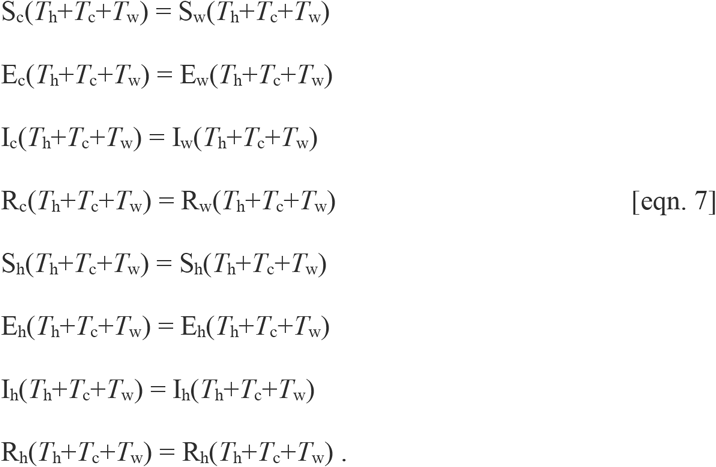

During this second commute phase (also of length *T*_c_), the infection dynamics for those commuting are given by [eqn. 3] above, and the infection dynamics for those still at home are given by [eqn. 4] above. At the end of the second commute phase, all individuals are back in the home environment (no longer subdivided) and the number of individuals of each type are

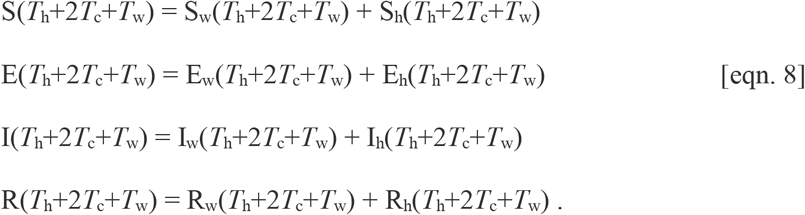

This ends the cycle for a single day; the next day starts the cycle again.

### Model Parameters

We used a fixed population size (*N*) of 3,000 individuals. We did not include births or deaths, or movement in and out of the population. These are reasonable assumptions given the scope of our simulations: a work population that is not hiring new employees and has few retirements or actual deaths over a few months. Because we assumed frequency-dependent transmission, the relative fraction of the population infected is the same regardless of population size.

Infection parameters were calculated as follows. We used 4.2 days as the latent period [37] and calculated the rate of moving from exposed to infected (σ) as the inverse of this: σ = 1/4.2 = 0.238 per day. We used 9.5 days as the infectious period (the estimated length of viral shedding for SARS-coV-2; [38]), and calculated the recovery rate (*γ*) as the inverse of this: *γ* = 1/9.5 = 0.105 per day. Transmission rate (*β*) was calculated based on the basic reproductive number, R_0_. We assumed a ‘baseline’ R_0_ (unmitigated; no behavioral changes like physical distancing) of 2.5 based on current estimates for SARS-coV-2 [39], although some estimates put R_0_ as high as 5.7 [37]. To quantify how behavioral changes to movement and contact affect transmission we defined effective reproduction numbers (R_e_) for each of the environments (home, work, commute). We assumed that stay-at-home measures to reduce pathogen spread in the community halved the rate of contacts at home (e.g., [40]), that is *R*_e-h_ = 0.5*R*_0_. We assumed that infection at work could be anywhere between current infection rates at home (*R*_e-w_ = 0.5*R*_0_) and unmitigated rates *(R*_e-w_ = *R*_0_). To facilitate interpretation of our results, we also describe infection at work in terms of the fraction increase in transmission compared to home, where 0 indicates transmission is the same at work and home, 0.5 indicates transmission at work is 50% higher than at home and 1 indicates transmission at work is 100% higher than at home (i.e., double). Finally, we assumed that infection while commuting spanned a broader range of possible values than either home or work. At one extreme, commuting by private transport effectively has no risk of transmission from others (*R*_e-c_ = 0). At the other extreme, commuting by crowded public transport can reduce feasible physical distancing (*R*_e-c_ = 2*R*_0_) both because individuals have a greater number of contacts while commuting and because these contacts potentially last for longer than normal. Transmission rates (*β*) were back-calculated from R_e_ values, based on rearranging the expression R_e_ = *β/γ* to *β* = *γ* R_e_. We assume that R_0_ and R_e_ values estimated for the general public apply to our population of University workers. If instead our population had lower transmission rates than the general public under stay-at-home measures, going back to work could lead to faster pathogen spread than we predict here.

### Simulations

Since our aim was to understand the relative importance of model parameters on infection dynamics (rather than try to forecast outcomes), we started each simulation with one individual infected (I(*t*=0)=1), zero exposed (E(*t*=0)=0), zero removed (R(*t*=0)=0), and the rest susceptible (S(*t*=0)=2,999). Each simulation was run until it reached equilibrium (where the fraction of the population in the *R* class did not change from one day to the next). We defined a baseline set of values for each parameter (see Table 2). Then we ran the following simulations that varied some parameters while holding others constant:

i. Varying transmission while at work (*β*_w_) and during the commutes (*β*_c_). We considered three scenarios that differed in the degree of risk of a commute to work and back. For low risk, we assumed low contact both during commutes and on campus (*R*_e-w_ = 0.5R_0_ = 1.25, equivalent to at home). For moderate risk, we assumed unmitigated contact during commute (*R*_e-c_ = R_0_ = 2.5, shared transport) and partial physical distancing at work (*R*_e-w_ = 0.75R_0_ = 1.875, intermediate between home and unmitigated). For high risk, we assumed elevated contact during commute (*R*_e-c_ = 2R_0_ = 5, crowded shared transport), and unmitigated contact at work (*R*_e-w_ = R_0_ = 2.5). These results are presented in Fig 2.
ii. Varying the fraction of the population commuting (*θ*) and fraction of the day spent on campus (*T*_w_). We considered eleven values of the fraction of the population commuting (*θ* = 0,0.1,…,0.9,1) and eleven values of the fraction of an 8-hour workday spent on campus (*T*_w_ = x(8/24) where x = 0, 0.1,…,0.9,1). These results are presented in Fig 3a.
iii. Varying the fraction of the population commuting (0) and fraction increase in transmission at work compared to home (*R*_e-w_). We considered eleven values of the fraction of the population commuting (0 = 0,0.1,…,0.9,1) and eleven values of the fraction increase in transmission at work compared to at home (*R*_e-w_ = (1+x)*R*_e-h_ where x = 0,0.1,…,0.9,1). These results are presented in Fig 3b.
iv. Varying the fraction of the day spent on campus (*T*_w_) and fraction increase in transmission at work compared to home (*R*_e-w_). We considered eleven values of the fraction of an 8-hour workday spent on campus (Tw = y(8/24) where y = 0,0.1,…,0.9,1), and eleven values of the fraction increase in transmission at work compared to at home (*R*_e-w_ = (1+x)*R*_e-h_ where x = 0,0.1,…,0.9,1). These results are presented in Fig 3c.

Movement model simulations were conducted in Matlab 2018b.

**[Fig 2.**
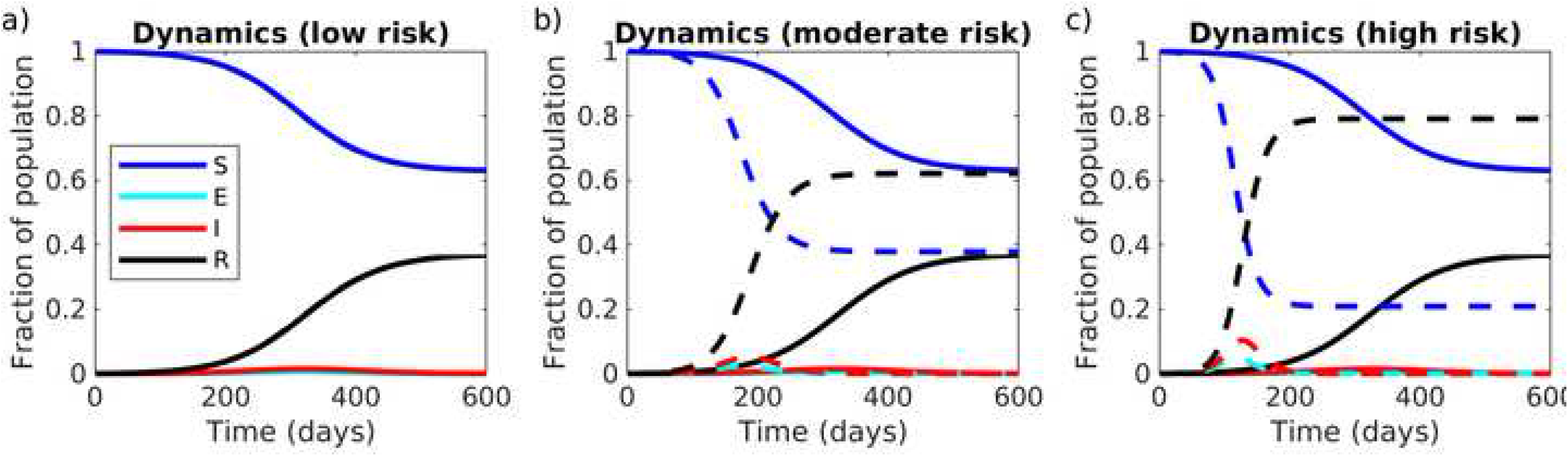
Movement model: varying the degree of physical distancing on campus and during commutes. The fraction of the population that is Susceptible (S), Exposed (E), Infected (I), and Removed (R), when all individuals either work from home (solid lines) or commute to work on campus (dashed lines), for different degrees of physical distancing both on campus and during the commute: (a) low risk: low contact during commute and on campus (*R*_e-c_ = *R*_e-w_ = 0.5R_0_ = 1.25, equivalent to at home), dashed and solid lines are identical, (b) moderate risk: unmitigated contact during commute (*R*_e-c_ = R_0_ = 2.5, shared transport) and partial physical distancing at work (*R*_e-w_ = 0.75R_0_ = 1.875, intermediate between home and unmitigated), (c) high risk: elevated contact during commute (*R*_e-c_ = 2R_0_ = 5, crowded shared transport), and unmitigated contact at work (*R*_e-w_ = R_0_ = 2.5).]

**[Fig 3.**
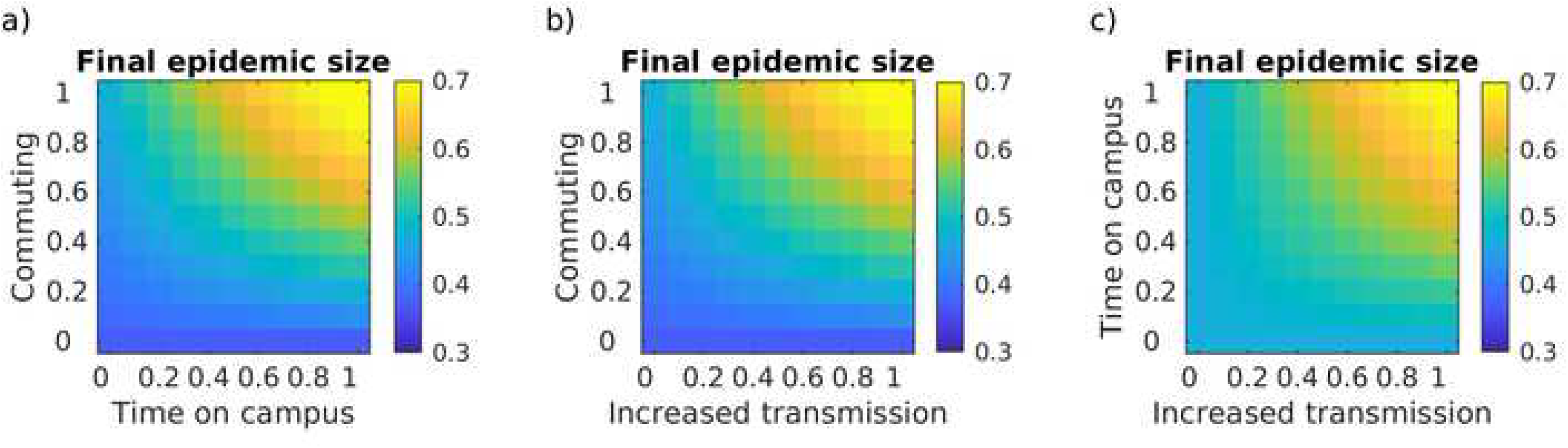
Movement model: limiting people, time and contact on campus. The final epidemic size (cumulative fraction of the population infected) as a function of (a) the fraction of an 8-hour workday spent on campus (x-axis) and the fraction of the population working on campus (y- axis) with no physical distancing, (b) the fraction increase in transmission while at work compared to at home (x-axis) and the fraction of the population working on campus (y-axis) with an 8-hour work day, (c) the fraction increase in transmission while at work compared to at home (x-axis) and the fraction of an 8- hour workday spent on campus (y-axis) with 100% of people on campus.]

### Sensitivity Analysis

Finally, we performed a sensitivity analysis to determine how sensitive the two model output metrics (final epidemic size, epidemic peak size) were to each of the model parameters, using a combination of Latin Hypercube Sampling (LHS) and Partial Rank Correlation Coefficients (PRCC). The LHS/PRCC sensitivity analysis is appropriate when the relationship between model output and each model parameter is monotonic and nonlinear [41]. For our model, this relationship was monotonic for all nine parameters considered (*N*, *T*_c_, *T*_w_, θ, σ, γ, *R*_e-c_, *R*_e-h_, and *R*_e-w_; S1-S2 Figs). The LHS/PRCC sensitivity analysis has two steps.

First, we used Latin Hypercube Sampling (LHS; [42]), a Monte Carlo approach, to generate sets of parameter value combinations from preset ranges of parameter values. LHS has a minimum required sample size (n) which is given by: *n* ≥ *k*+1 or *n* ≥ *k*(4/3) where *k* is the number of parameters included in the LHS [43], nine for our analysis. We chose the number of samples (see below) to meet these criteria. Each of the nine model parameters considered was sampled from a uniform probability density function based on the ranges given in Table 2. The model was run for each parameter value set, and the final epidemic size (cumulative fraction of the population infected, in the long-term) and epidemic peak size (maximum fraction of the population infected at any time) were both saved as output metrics.

Second, we measured the sensitivity of the output metrics to each parameter using Spearman Partial Rank Correlation Coefficients (PRCC). To determine how many samples of each parameter was needed to generate stable PRCC value, we calculated PRCC value for an increasing number of samples (S3 Fig) and noted that the PRCC values were relatively stable past 1000 samples. Thus, we used 1000 samples of each parameter value for our final PRCC analysis. A positive PRCC value indicates that increasing the value of that parameter increases the output metric while a negative PRCC value indicates that increasing the value of that parameter decreases the output metric. PRCC values that were not significant at the 0.05 level are marked with ‘ns’ in Fig 4 (not corrected for multiple comparisons).

Finally, we used a z-test to rank significant model parameters in terms of their relative importance, since larger PRCC values do not always indicate more important parameters [41]. For our results (Fig 4), model output sensitivity was indeed given by the size of PRCC values.

**[Fig 4.**
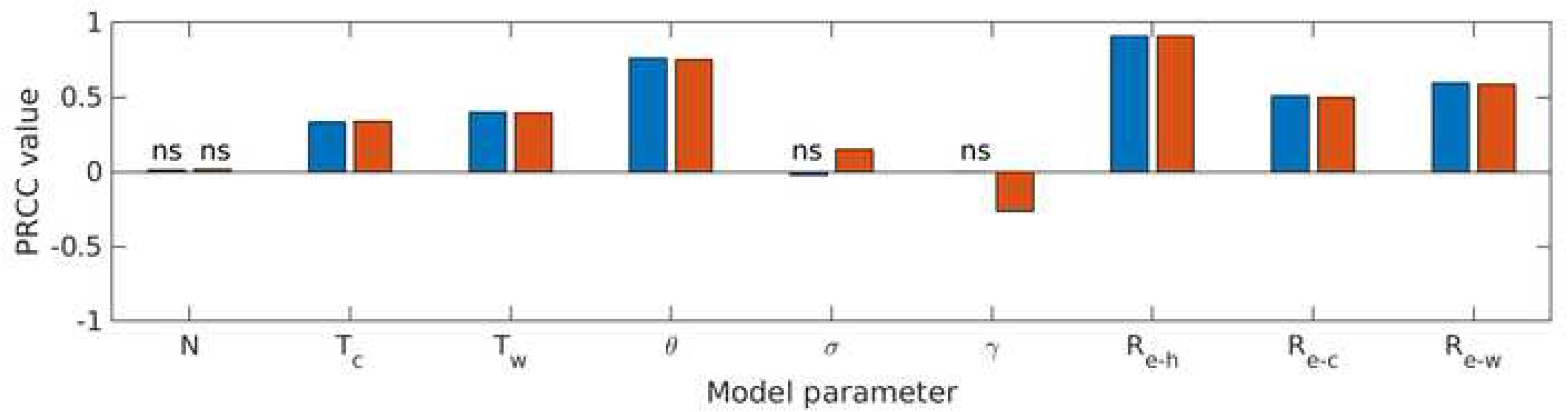
Movement model: sensitivity analysis. The partial rank correlation coefficient (PRCC) values for each model parameter (Table 2) for the final epidemic size metric (blue bars) and the epidemic peak size metric (orange bars). Positive values indicate parameters that increase epidemic size as they are increased (negative values indicate parameters that decrease epidemic size as they are increased). Cases where the relationship between the parameter and model output metric was not significant are indicated with ‘ns’.]

### Methods: network model

Our second model explores infection dynamics as individuals work on campus either in both office and lab spaces or just in lab spaces. We created a network map of all the individuals housed in the Ecology building on the St. Paul campus of the University of Minnesota. We created our dataset by merging information on the office and lab room assignments for each individual with an office or lab in the building. (The methods for collection and analyses of these data were reviewed by the University of Minnesota’s Institutional Review Board and were determined not to be human subjects research.) Work in the Ecology building is structured by two primary space types, laboratories that can include one to three research groups, each associated with a single faculty member, and offices which can be single-occupancy or shared. Office space is generally shared by groups of graduate students and postdoctoral scholars, often from different lab groups. Because undergraduates are generally not permitted to work on campus during the resumption of research, we included faculty, staff, postdocs, and graduate students, but excluded all undergraduates from this visualization.

We considered two types of bipartite networks: shared office space and shared lab space. Individuals sharing an office or a lab all had an edge with that location node. We then consider the one-mode projection of each network, creating a weighted unipartite network connecting individuals according to their shared spaces. The binary representation of these networks was used to create static network visualizations of connections among individuals using the igraph, tidygraph, and ggraph libraries in R [44-46], shown in Fig 5a and b. Animations of disease progression through the networks were produced using the gganimate library in R (S5-S6 Figs; [47]). For each network, we computed the distribution of (finite) shortest paths between each pair of nodes (Fig 5c) and for each distinct component of the networks, we noted its size (number of nodes), diameter (longest shortest path), and mean path length (average shortest path length; S7 Fig).

**[Fig 5.**
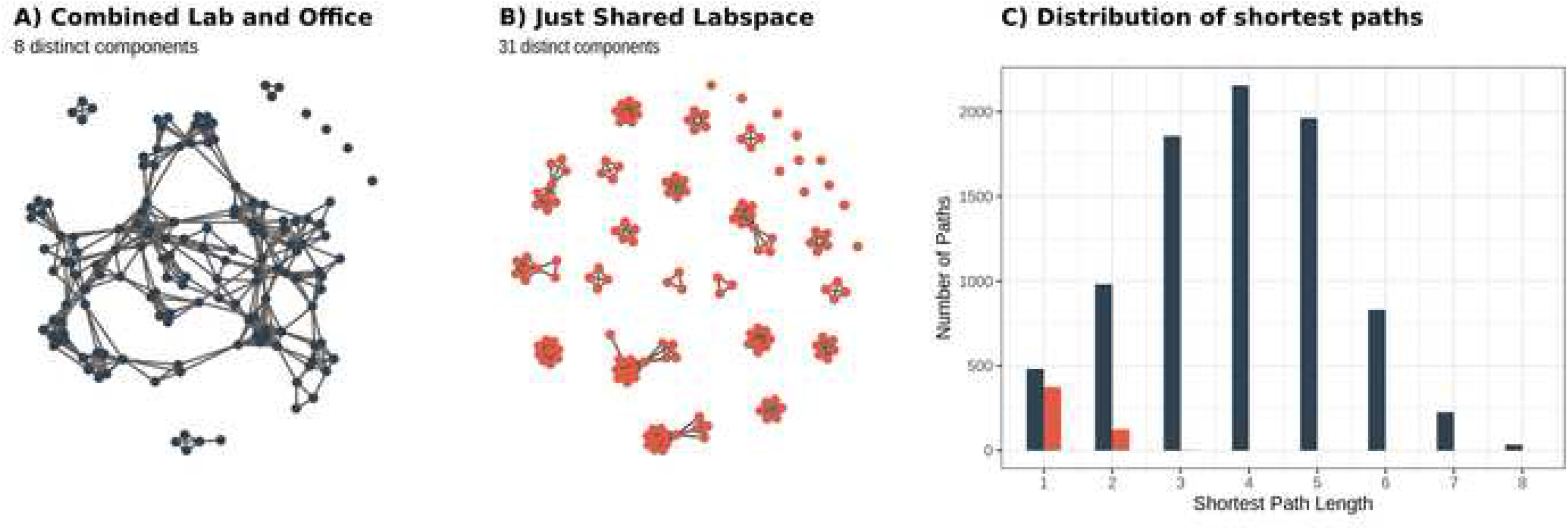
Network model structure. Space-sharing, or ‘contacts’ (edges) are shown among all individuals (nodes) for two scenarios: (a) when individuals at work share either office or lab spaces, or (b) when individuals only used shared lab space and not shared offices (e.g., bench work is done on campus while office work is done at home). (c) Histograms showing the distribution of shortest paths between all connected pairs of individuals. Importantly, though all shortest paths between nodes in the network containing only links of shared lab spaces are less than or equal to three, the vast majority (approximately 95%) of pairwise combinations of individuals actually have no chain of interactions connecting them. In contrast, the combined network contains a component consisting of almost 90% of individuals in the network, corresponding to nearly 80% of all pairs of individuals having a chain of interactions connecting them.]

For the network simulations, we used an SEIR model framework, starting with a randomly selected index case to serve as the first infected individual in an entirely susceptible population (Fig 6). Simulations proceeded in discrete time. At each time step, individuals who had been exposed to the virus transitioned into the infectious class (E→I) based on the result of a Bernoulli trial using the disease progression rate as the probability of success. Likewise, currently infectious individuals were removed (i.e., either recovered and immune or deceased; I→R) based on the result of a Bernoulli trial using the recovery rate as the probability of success. Finally, one Bernoulli trial using the transmission rate as probability of success was conducted for each edge connecting a susceptible individual to an infectious one. Susceptible individuals became exposed (S→E) if at least one such trial resulted in success. At the end of each simulation, we took note of the epidemic peak size, the final epidemic size, and the time needed to read the epidemic peak (Fig 7). We evaluated the sensitivity of these results by comparing them to simulations run on randomized versions of these empirical networks (S8 Supporting Information). Network analysis and simulations were conducted in R (Version 3.6.3).

**[Fig 6.**
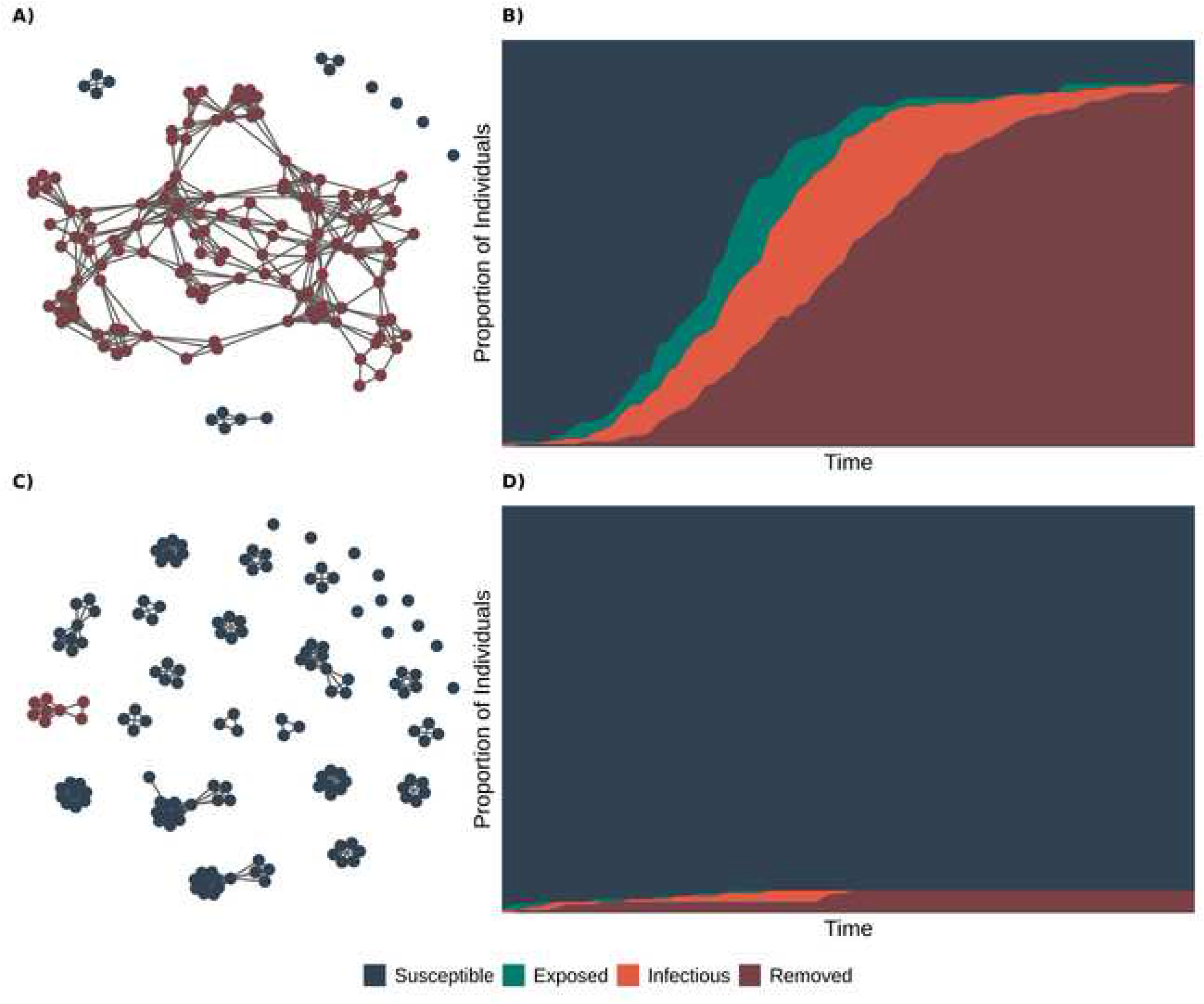
Network model simulations. Final disease status of members of networks based on use of (a) both shared lab and office space and (c) only shared lab space, following a simulated epidemic with susceptible individuals in blue, exposed individuals in green, infectious individuals in orange, and removed individuals in red. (b, d) the cumulative number of susceptible, infectious, and removed individuals over time for each network simulation.]

**[Fig 7.**
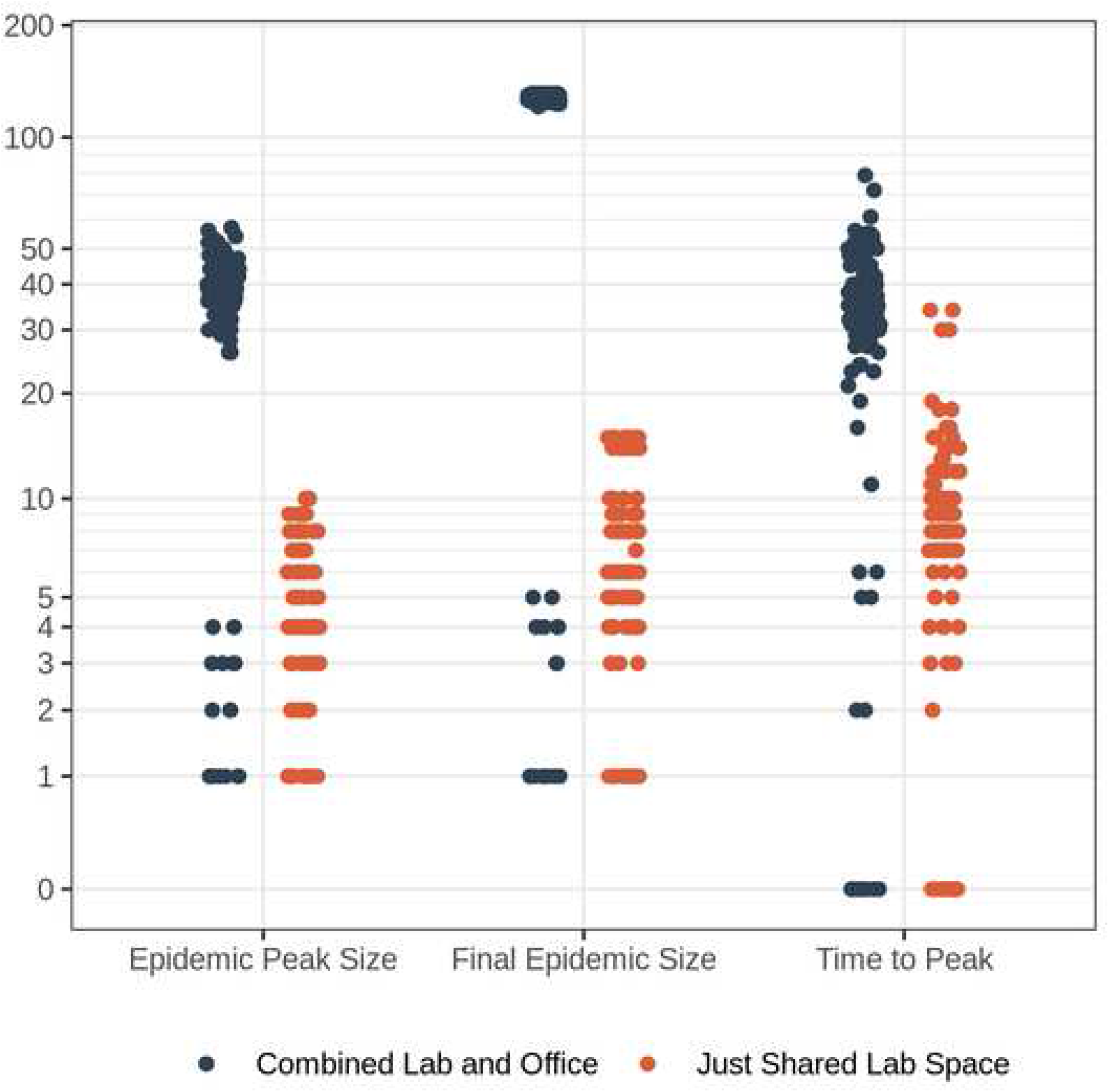
Network model simulations. Outcome of 100 infection simulations on networks: the maximum peak number of individuals infected at any one time (epidemic peak size), total number of individuals infected (final epidemic size), and time until peak number of individuals infected for simulations of pathogen spread on networks based on use of both shared lab and office space (blue) and only shared lab space (orange).]

## Results

### Movement model

Whether returning to work on campus affects the epidemic outcomes (measured as final epidemic size and epidemic peak size) depends critically on the degree of physical distancing maintained both on campus and during the commute between home and campus (Fig 2). If the current degree of physical distancing that is achieved while working from home can be maintained while on campus, then working from campus will not speed up infection dynamics compared to working from home (Fig 2a). However, if physical distancing on campus or during the commute is less successful than current physical distancing at home, then returning to work on campus will both increase the epidemic peak size in the short-term and increase the final epidemic size in the long term (Fig 2b-c). When physical distancing cannot be maintained on campus or during the commute, then infection dynamics can be kept slower by limiting the fraction of workers on campus and the amount of time workers are on campus (Fig 3a).

Intriguingly the three strategies we considered (limiting contact, people, or time on campus) are interchangeable with approximately equivalent effects on both the long-term metric, final epidemic size (Fig 3) and the short-term metric, epidemic peak size (S4 Fig). That is, in situations where one of these strategies cannot be fully implemented, a different strategy can be used in its stead. For example, if individuals need to be on campus for an extended period of time to run an experiment (thus limiting time on campus is not a feasible strategy), this can be compensated for by limiting the number of other individuals on campus at the same time. However, of the three strategies, reducing the fraction of the population on campus had a bigger impact than reducing either time or contact on campus, due to the effect of commuting to and from campus. Regardless of time or physical distancing on campus, more people working on campus requires that more people commute. Thus, if commuting substantially increases transmission risk compared to staying at home (i.e., any form of shared transport vs. commuting alone), reducing the number of people commuting will be a more effective strategy than reducing either time or contact while on campus.

The sensitivity analysis revealed that both model metrics (final epidemic size, epidemic peak size) were most sensitive to transmission at home (*R*_e-h_), since most of the day is spent in that environment, as well as the fraction of the population commuting (*θ*) to campus (Fig 4). Transmission on campus *(R*_e-w_) and transmission during commutes *(R*_e-c_) were the next most influential; the first because most time during the workday is spent on campus and the second because we allowed transmission to vary across a wider range during commuting than on campus. The time spent on campus (*T*_w_) and time commuting (*T*_c_) were somewhat influential. For each of these parameters, increasing the parameter value increased the final and peak epidemic sizes. Finally, population size *(N)* did not significantly affect either metric (but would be critical for the total number of individuals infected). The rate of moving from exposed to infected (σ) and the recovery rate (γ) did not significantly affect final epidemic size, but both had a minor effect on epidemic peak size: increasing σ (i.e., a shorter latent period) increased epidemic peak size, while increasing *γ* (i.e., a shorter infectious period) decreased epidemic peak size.

### Network model

The mixing of researchers from different labs in shared office spaces had a substantial impact on the modularity of the resulting network. In particular, when people do not use shared office spaces (i.e., work from home if they share an office), but work on campus only in labs, the network is far more modular, with smaller, more densely connected groups and few connections among groups (Fig 5b, S7 Fig). In this case, most individuals are directly connected to all other members of their group (i.e., “shortest path” of one, Fig 5c); however the absence of connections between groups means that, on average, an infected individual lacks a path of connections to 95% of the rest of the population. In contrast, when individuals share both lab and office space, the connectedness of the network is relatively high because students, staff, and postdocs that share offices are often from different labs. For this combined case, most individuals are four or fewer connections from one another (Fig 5c) and the largest component contains nearly 90% of all individuals in the network (Fig 5a, S7 Fig). Thus, in the case where an infected individual (presumably pre-symptomatic or asymptomatic) came to work, the combined lab and office network has the potential for greater disease incidence than in the lab-only network, where the infection could be constrained to a single lab (Figs 6-7, S5-S6 Figs). In general, when compared to the combined network, the lab-only network had outbreaks that were less explosive (i.e., had less variance and a lower mean number of individuals infected at any one time), fewer individuals infected overall, and a shorter time until the peak number of infectious individuals (Fig 7).

## Discussion

Movement and contact behaviors are key drivers of the spread of pathogens like SARS-CoV-2, and not all movement and contacts have the same impact on pathogen spread. However, basic compartmental models used to describe SARS-CoV-2 dynamics assume all individuals move and contact each other at random (i.e., populations are well-mixed). Our models show how explicitly accounting for movement, space use in a building, and contact behaviors can provide a more nuanced understanding of relative risk. Our movement model, capturing the predictable movement between home and work/campus environments, shows that reducing the number of people, rate of contact, and amount of time spent on campus are all equivalently effective strategies for slowing pathogen spread. However, if commutes specifically increase transmission risk (i.e., shared transport), reducing the number of people on campus is the most effective strategy to reduce the infection spread rate. We also considered heterogeneity in contact behavior once at the workplace; our network model captures the regular interactions among workers in shared workspaces on campus and shows that restricting building use to lab spaces (rather than lab and office space) may reduce pathogen spread. Our results provide a number of tools to distinguish among different movement and contact patterns at the scale of individuals and workplace communities.

A number of future directions could be explored, by changing some of our simplifying assumptions. First, staying within the broad structure of our model, alternative spatiotemporal strategies could be explored including: structured work weeks (e.g., four days on-campus and ten off; [48]), or further compartmentalizing time (e.g., sequential work shifts) or space (e.g., different buildings on campus). For instance, if evidence suggests that infection can occur through air circulation within buildings [49], these models could be altered to account for connections arising from shared ventilation systems. These models also could be modified to account for movement and contact behavior that explicitly depends on infection status [50]; e.g., splitting infected individuals into infectious but asymptomatic (who still potentially commute to work) and symptomatic individuals (who stay home) or building dynamic networks where contact behavior can change in response to infectious status (e.g. infectious and symptomatic individuals reducing their interpersonal contacts either through staying home or altering their behavior at work). Second, one could expand the scale of the model. This could be done foremost by combining the movement model (movement between work, commute, campus) with the network model (movement while on campus). Further expansions could consider both larger scales (linking in regional patterns) as well as smaller ones (allowing contacts within buildings to vary over time). For instance, integrating local models such as ours with regional variation in infection rates and degree of community social mixing [11,51,52] could further inform recommendations. Third, as data accumulate on transmission dynamics and individual susceptibility, we can alter specific players or interactions in the model. For instance, while the virus can survive on surfaces [53], most transmission appears to be aerosolized, mediated by extended person-to-person interactions in close spaces [54-58], so masking and minimizing temporal and spatial overlap of workers in shared spaces is key [59-61]. In addition, susceptibility and thus local demographic data can provide additional layers of tailored recommendations [62].

Our findings mesh with concepts in the broader movement and disease ecology literature. Within movement ecology, there has long been a distinction between random/undirected movement like dispersal versus predictable movements like diel and seasonal migration [63]. Human movement between home and work is often a predictable and daily occurrence and thus is better viewed from the lens of predictable migratory movements (as we do here) rather than random dispersive ones (as implicit in basic compartmental models). Moving predictably between two environments does not always increase infection (either for individuals or at the population level) compared to remaining in a single location; the relative transmission in each environment is critical [22]. We find that transmission risk during a commute is key to infection dynamics when considering the impact of movement between home and work, paralleling recent work calling for the explicit consideration of how transient phases of movement affect infection dynamics [24] and theory showing that infection dynamics during transit can have a similar impact to dynamics in the second environment [64].

There are important insights that emerge from our movement and contact-network models that can guide policy. For example, basic disease models assume random movement and equal probability of contact, whereas many hosts, including humans, move in directed ways and in very structured social networks. For this reason, disease mitigation policies will likely be more effective when they consider disease risk in a more holistic way that integrates risk across the various components of a person’s daily movement. For example, in settings where many people commute by mass transit (e.g., New York City), the efficacy of workplace safety protocols may be overwhelmed by transmission during daily commutes rather than contacts at work. Careful examination of social network patterns could also help guide policy to provide intermediate scenarios between business as usual and complete lock down. For example, in our case-study contact rates and potential disease spread were significantly reduced when people’s contacts at the workplace were restricted to single lab groups, as opposed to linking separate lab and office networks. These findings are consistent with emerging calls to reduce COVID-19 spread by creating “learning pods” and “social bubbles” of interacting children and adults as schools and workplaces re-open [65,66].

Even so, the protective effects of heterogeneity in contact structure should not be overemphasized for decision making. First, although the threshold for herd immunity can be lower in heterogeneous networks [67], making outbreaks less likely, outbreaks that do occur can also be more explosive [25]. Second, because SARS-CoV-2 spread appears to be primarily by aerosolized transmission, the potential contact behaviors needed for transmission are more ubiquitous than for pathogens with more specific transmission modes (e.g., sexually transmitted diseases like HIV/AIDS). Importantly, the networks presented here consider only the room in which an employee works (their office or lab space), explicitly omitting broader workplace considerations like air flow, shared surfaces, entry points, etc., these additional points must be addressed in conjunction with thinking about explicit contact behavior when forming a public health strategy. Lastly, these static networks are a simplification of an inherently dynamic process of movement, contact, and infection. Using a time-ordered or dynamic network approach could provide better insights to actual duration of exposures and sickness-induced behavioral changes [68].

## Conclusions

Human movement and contact behaviors are critical for the spread of pathogens like SARS-CoV-2, yet are rarely addressed explicitly in the current conversations about decision-making in the face of relaxing Stay at Home orders. Here we have drawn on movement and network models to demonstrate the effect of these behaviors. First, we have shown that regular movement between two ‘environments’ (i.e., work and home) does not inherently increase infection spread the way random dispersive movements might. Rather the outcome depends on the relative degree of transmission (e.g., degree of physical distancing) in each environment. Second, we have shown that different contact patterns (e.g., space usage) within the work environment could lead to different outcomes in terms of SARS-CoV-2 spread. In sum, we advocate for using an understanding of movement and contact patterns as an adjunctive approach (alongside widespread testing, contact tracing, vaccine development and other tools) to mitigate the effects of SARS-CoV-2 and COVID-19, particularly when considering return to work environments.

## Data Availability

This manuscript is a theoretical study and does not contain health-related data. The model code and output will be made publicly available via GitHub and Data Dryad when the paper is published in a journal.

## Acknowledgements

We thank William Harcombe for helpful feedback and discussion, and Valery Forbes, David Greenstein and Daniel Stanton for encouragement.

## Data Availability

Model code and simulation output for the movement model is available on Data Dryad [69]. All Code to generate the network model figures and animations is available on Github (https://github.com/whit1951/EEBCovid).

---

SUPPORTING INFORMATION FIGURE CAPTIONS

**S1 Fig.** Movement model monotonicity plots. The relationship between each of the nine model parameters (x-axis) and the model output, final epidemic size (y-axis) for (a) population size (*N*); (b) fraction of a 24-hour day spent commuting each way for those that commute to campus (*T*_c_); (c) fraction of a 24-hour day spent on campus for those commuting (*T*_w_); (d) fraction of the campus population commuting to work on campus (*θ*); (e) recovery rate (*γ*); (f) effective reproductive number while at home (*R*_e-h_); (g) effective reproductive number while commuting between work and campus (*R*_e-c_); and (h) effective reproductive number while at work on campus (*R*_e-w_).

**S2 Fig.** Movement model monotonicity plots. The relationship between each of the nine model parameters (x-axis) and the model output, epidemic peak size (y-axis) for (a) population size (*N*); (b) fraction of a 24-hour day spent commuting each way for those that commute to campus (*T*_c_); (c) fraction of a 24-hour day spent on campus for those commuting (*T*_w_); (d) fraction of the campus population commuting to work on campus (□); (e) recovery rate (*γ*); (F) effective reproductive number while at home (*R*_e-h_); (g) effective reproductive number while commuting between work and campus (*R*_e-c_); and (h) effective reproductive number while at work on campus (*R*_e-w_).

**S3 Fig.** Movement model sample numbers. Absolute value of PRCC for the final epidemic size model output and each of the nine model parameters (*N*, *T*_c_, *T*_w_, □, σ, *γ*, *R*_e-h_, *R*_e-c_, *R*_e-w_) as a function of different numbers of LHS samples generated. The results seem to stabilize after about 1000 samples.

**S4 Fig.** Movement model: limiting people, time and contact on campus. The epidemic peak size (maximum fraction of the population infected) as a function of (a) the fraction of an 8-hour workday spent on campus (x-axis) and the fraction of the population working on campus (y- axis) with no physical distancing, (b) the fraction increase in transmission while at work compared to at home (x-axis) and the fraction of the population working on campus (y-axis) with an 8-hour work day, (c) the fraction increase in transmission while at work compared to at home (x-axis) and the fraction of an 8- hour workday spent on campus (y-axis) with 100% of people on campus.

**S5 Fig.** Network model simulations. Simulations of pathogen spread across networks based on use of both shared office and lab space.

**S6 Fig.** Network model simulations. Simulations of pathogen spread across networks based on use of only shared lab space.

**S7 Fig.** Component-wise network structural metrics. Measures of the size (number of individuals), diameter (longest shortest path between two individuals), and mean path length (average shortest path length between individuals) for each distinct component of networks presented in Fig 5a,b. The combined lab and office network (blue points) has 8 distinct components (8 points for each metric), while the shared lab space network contains 31 distinct components (31 points for each metric).

**S8.** Supporting Information. Sensitivity analysis of the network model and results.

